# General SIR model for visible and hidden epidemic dynamics

**DOI:** 10.1101/2024.12.17.24319157

**Authors:** Igor Nesteruk

## Abstract

To simulate hidden epidemic dynamics connected with asymptomatic and unregistered patients, a new general SIR model was proposed. For some cases, the analytical solutions of the set of 5 differential equations were found which allow simplifying the parameter identification procedure. Two waves of the pertussis epidemic in England in 2023 and 2024 were simulated with the assumption of zero hidden cases. The accumulated and daily numbers of cases and the duration of the second wave were predicted with rather high accuracy. If the trend will not change, the monthly figure of 9 new pertussis cases (as it was in January-February 2023) can be achieved only in May 2025. The proposed approach can be recommended both for simulations and predictions of different epidemics.

## 1. Introduction

Asymptomatic and unregistered cases are characteristic of almost all infectious diseases, in particular, SARS-CoV-2 [1–6] and pertussis [7] are no exception. The percentage of asymptomatic patients can be age dependent and lead to huge differences in registered numbers of cases for countries with young and old population [8–10]. Some theoretical estimations of the visibility coefficient *β* - the ratio of real infections to the registered ones can be found in [9, 11–13]. In this study we will use the concepts of the classical SIR (susceptible-infectious-removed) model [14–20], its generalization for simulations of different epidemic waves [20–22] and procedures of parameter identification [22–23]. Numerous improvements of SIR model (see e.g., [24–28]) do not take into account the visibility coefficient.

The obtained theoretical results will be applied for simulations of the pertussis (whooping cough) epidemic in England in 2023 and 2024 [29]. This disease increases the risk of infant fatality and became a serious problem in many countries including the developed ones [27, 29]. Numerical differentiation of the monthly numbers of new cases revealed two waves of the epidemic in England (before and after November 2023) [30]. Due to the absence of necessary amount of observations, SIR simulations in [30] were performed in [30] only for the first wave. In this study we will use the new approach for simulation of both waves of the pertussis epidemic and compare the predictions with the recent statistical data.

## 2. Differential equations and initial conditions

For every epidemic wave *i*, let us divide the compartment of infectious persons *I(t)* (*t* is time) into visible (registered) and hidden (invisible/asymptomatic and unregistered) parts *I* = *I* ^(*v*)^ + *I* ^(*h*)^ and suppose that these persons are appearing according to the visibility coefficient *β* ≥ 1and removing with rates 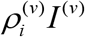 and 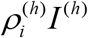. Then the general SIR model [20–22] takes the following form:

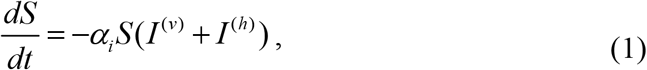

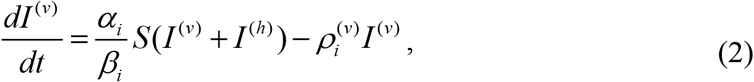

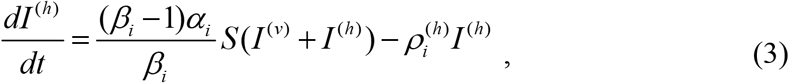

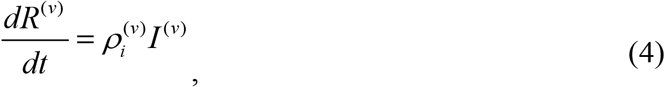

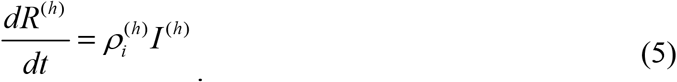

The compartment of removed persons *R(t)* is also divided into visible (registered) and hidden parts *R* = *R*^(*v*)^ + *R*^(*h*)^. Infection and removal rates (*α*_*i*_,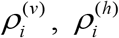) and the visibility coefficient *β*_*i*_ are supposed to be constant for every epidemic wave, i.e. for the time periods: 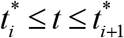, *i* = 1, 2,3,…. Summarizing eqs. (1)–(5) yields zero value of the derivative *d* (*S* + *I* ^(*v*)^ + *I* ^(*h*)^ + *R*^(*v*)^ + *R*^(*h*)^) / *dt*. Then the sum:

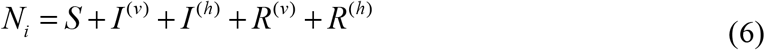

must be constant for every epidemic wave. We will consider the value *N*_*i*_ to be an unknown parameter of the model corresponding to the *i-th* wave, which is not equal to the known volume of population and must be estimated by observations. There is no need to assume that before the outbreak all people are susceptible, since many of them are protected by their immunity, distance, lockdowns, etc. Thus, we will not reduce the problem to a 4-dimensional one. It means that the solution can be obtained by numerical integration of the set of 5 differential equations (1)–(5). Nevertheless, there are some separate cases, when analytical solutions are possible (see next Section).

Taking into account (6), the initial conditions for the set of equations (1)–(5) at the beginning of every epidemic wave 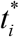 can be written as follows:

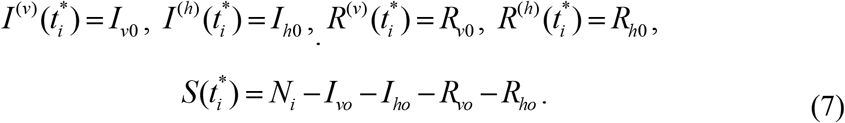

If at moment 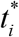 all previously infected persons are removed, we can take into account only cases starting to appear during *i-th* wave and use the initial conditions:

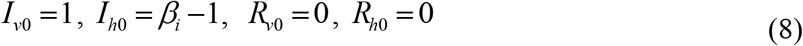

### 3. Examples of analytical solutions

Let us introduce the functions corresponding to the accumulated numbers of visible and hidden cases :

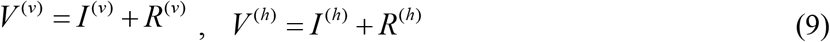

Then it follows from (2)–(5) that

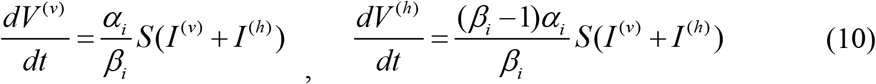

Dividing (10) by (1) yeilds:

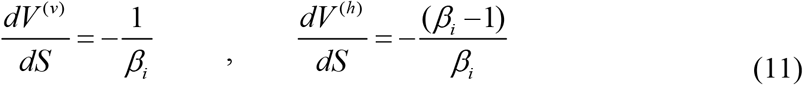

and simple linear solutions taking into account initial conditions (7):

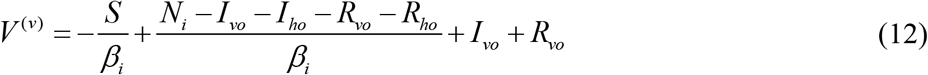

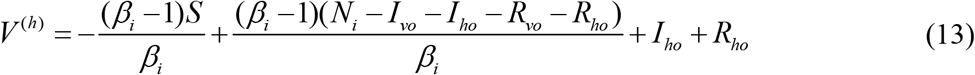

Eqs. (12) and (13) allow obtaining simple linear relationship:

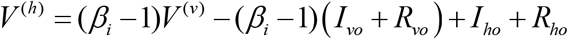

which demonstrates that the ratio of total accumulated cases *V* = *V* ^(*v*)^ +*V* ^(*h*)^ to the registered ones:

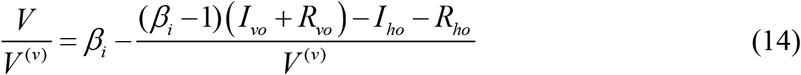

is not constant even for initial conditions (8) and equals *β*_*i*_ only approximately at large *V* ^(*v*)^ numbers. Eq. (14) limits the accuracy of the approach used in [11–13].

Introducing

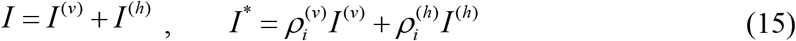

summarizing (2) and (3) and dividing by (1) yield the following differential equation:

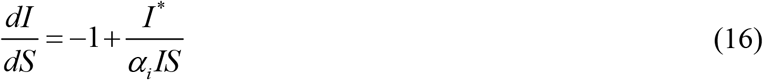

In 3 separate cases:

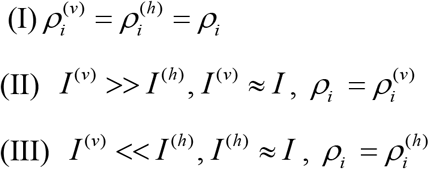

eq. (16) simplifies and has an analitycal solution taking into account the initial conditions (7):

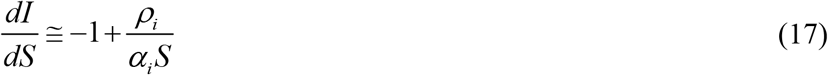

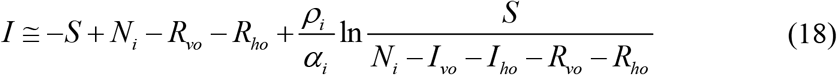

Eqs. (17) and 18 exact in the case (I) and approximate in cases (II) and (III).

Putting equation (18) into (1) and integration yield:

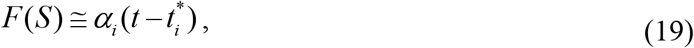

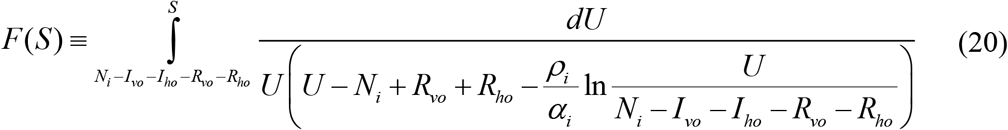

It follows from (1), (2) and (15) that:

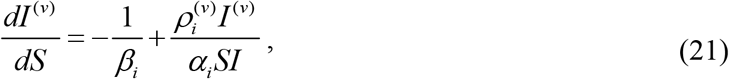

Taking into account that

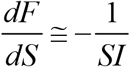

(see (18) and (20)), the solution of the non-homogenous linear equation (21) satisfying the first initial condition (7) can be written as follows:

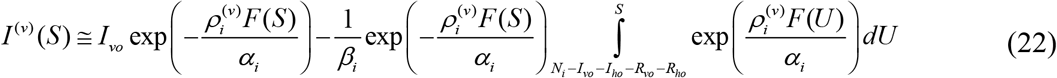

With the use of (9) and (15) it is possile to express other functions as follows:

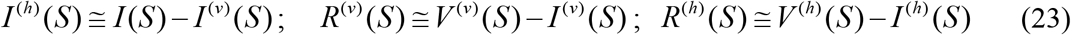

Then for every value of S, all unknown functions can be calculated with the use of. (12), (13), (18), (22) and (23). Corresponding moments of time can be found with the use of (19), (20). Thus, eqs. (12)–(13), (18)–(20), (22), (23) yield an approximate analytical solution of the set of differential eqs. (1)–(5) with the initial conditions (7). In the case (I) and when *I* ^(*v*)^ = *I*, this solution is exact. For *I* ^(*v*)^ = *I*, there is no need in (22), (23) and corresponding formulas obtained in [20–22] are also valid.

## 4. Examples of parameter identifications and predictions

The analytical solution simplifies the procedure of identification of unknown parameters, since there is no need in numerical integration of differential equations (1)–(5). It particular, having the set of accumulated cases 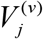 registered at moments *t*_*j*_, we can calculate corresponding values *S*_*j*_ with the use of the linear equation (12) for any values of unknown constant parameters appearing in eqs. (1)–(7). Then eq. (20) allows calculating values *F*_*j*_ *=F(S*_*j*_). Due to the linear relationship (19), standard linear regression formulas [31] can be used to calculate the correlation coefficient *r* and values of parameters *α*_*i*_ and 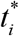. The optimal values of model parameters (providing the best fitting between the theoretical *V* ^(*v*)^ (*t*) curves and the results of observations 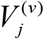) correspond to the maximum value of the correlation coefficient *r*. Thus, the parameter identification problem can be reduced to the problem of searching the maximum of complicated but analytical function *r*. For *I* ^(*v*)^ = *I* (a completely visible epidemic), such approach was successfully used to simulate and predict the dynamics of mysterious children disease [23], COVID-19 pandemic [20–22] and the pertussis epidemic in England [30].

Let us illustrate the parameter identification procedure for two waves of the pertussis epidemic in England in 2023 and 2024 discussed in [30]. The accumulated confirmed numbers of cases 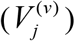 and corresponding moments of time *t*_*j*_ are listed in Table 1 according to the official site of UK government ([29], version available on 10 August 2024, the last 4 figures were taken from 10 December 2024 version). The values 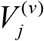 were used to calculate approximate daily numbers of new cases *dV/dt* at moments *t*_*j*_ according to the formula (1) from [30] (see the last column in Table 1).

**Table 1.** Accumulated numbers of confirmed pertussis cases in England in 2023 and 2024 and estimations of the average daily numbers of visible cases.

| Months,<br>starting with<br>January 2023 | Days, starting<br>with 1 January<br>2023, $t_j$ | Accumulated<br>numbers of<br>visible cases,<br>$V_j^{(v)}$, [29] | Calculated daily<br>numbers of new<br>visible cases at<br>moments $t_j$ $dV/dt$, eq.<br>(1) in [30] |
| --- | --- | --- | --- |
| 1 | 31 | 9 | - |
| 2 | 59 | 18 | 0.35426 |
| 3 | 90 | 30 | 0.52688 |
| 4 | 120 | 50 | 0.86559 |
| 5 | 151 | 83 | 1.41559 |
| 6 | 181 | 136 | 2.04462 |
| 7 | 212 | 208 | 2.66129 |
| 8 | 243 | 301 | 3.20 |
| 9 | 273 | 403 | 3.34516 |
| 10 | 304 | 505 | 3.47849 |
| 11 | 334 | 615 | 5.75269 |
| 12 | 365 | 858 | 12.87096 |
| 13 | 396 | 1413 | 24.79644 |
| 14 | 425 | 2332 | 38.86096 |
| 15 | 456 | 3759 | 58.23280 |
| 16 | 486 | 5872 | 84.44247 |
| 17 | 517 | 8924 | 89.67581 |
| 18 | 547 | 11351 | 67.65968 |
| 19 | 578 | 13038 | 44.27419 |
| 20 | 609 | 14096 | 28.79785 |
| 21 | 639 | 14800 | 19.94301 |
| 22 | 670 | 15309 | - |

Since the general problem contains 10 unknown parameters, their identification needs high performance computing even for the analytical solution (12)–(13), (18)–(20), (22), (23). When this solution is approximate, the optimal values of parameters will contain discrepancies, which can reduce the accuracy of predictions. For our example, let us take the case of exact solution *I* ^(*v*)^ = *I* (*β* = 1) and assume that at the beginning of every new epidemic wave all infectious persons from the previous waves are removed. Then we can use initial conditions (8), provide simulations and then add cases accumulated at moments when the monthly numbers of visible cases started to increase.

For every wave we will have only four unknown parameters *N*_*i*_, *α*_*i*_, 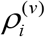 and 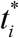. Due to linear relation (19) and using linear regression, only two of these parameters are independent.

The first and second epidemic waves were simulated with the use of 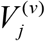 and *t*_*j*_ corresponding to *j*=3-10 and *j*=11-18, respectively. The optimal values of parameters (corresponding to the maximum of correlation coefficients r=0.999605701810268 (0.999822556136920)) are:

N_i_=20733.2400000011 (3657890.47292358);

*α*_*i*_ =2.07680793647070e-05 (2.11266650706657e-06) [day]^-1^;

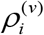 =0.417437564507461 (7.7088126606047) [day]^-1^;

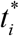 =57.6424779286801 (333.429529143536) days;

(figures in brackets correspond to the second wave). It should be noted that these optimal values are very different for the first and second waves and differ from the figures in [30] for the simulation of the first wave using observations with *j*=1-10.

Accumulated numbers of visible pertussis cases (blue cuves, the first eq. (9)); the average daily numbers of new cases (black curves, the first eq. (10)); numbers of infectious persons (red curves, eq. (18)). “Circles” represent the confirmed numbers of cases 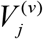 taken for identification of parameters of the first (*j*=3-10) and second (*j*=11-18) waves; blue “crosses” – all confirmed numbers of cases 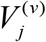 listed in Table 1; black “crosses” – results of calculations of approximate daily numbers of new cases at moments *t*_*j*_ listed in the last column of Table 1.

Using these optimal values in the analytical solution (10), (12), (18)–(20), (22), (23) yielded the theoretical curves shown in Fig. 1 (solid and dashed for the first and the second wave, respectively). The predicted values (see blue and black “crosses” for *j*=19-22, July-October 2024) are in good agreement with the theoretical blue and black curves. To estimate the accuracy of 4-month prediction, let us take the accumulated number of visible cases 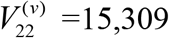 corresponding to the end of October 2024 (see Table 1) and compare with the theoretical value *V* ^(*v*)^ | _*t* = 670_ = 17,430 corresponding to the blue dashed line in Fig. 1. After adding 505 cases accumulated at *t*_*10*_ and extracted for the simulation of the second wave (compare blue “crosses” and “circles” in Fig.1), we obtain the accuracy (17,935-15,309)/15,309 around 17%. The average daily numbers of new cases will be less than 1.0 only in March 2025. If trend will not change, the monthly figure of 9 new cases (as it was before starting the first wave, see Table 1) can be achieved only in May 2025.

**Fig 1.**
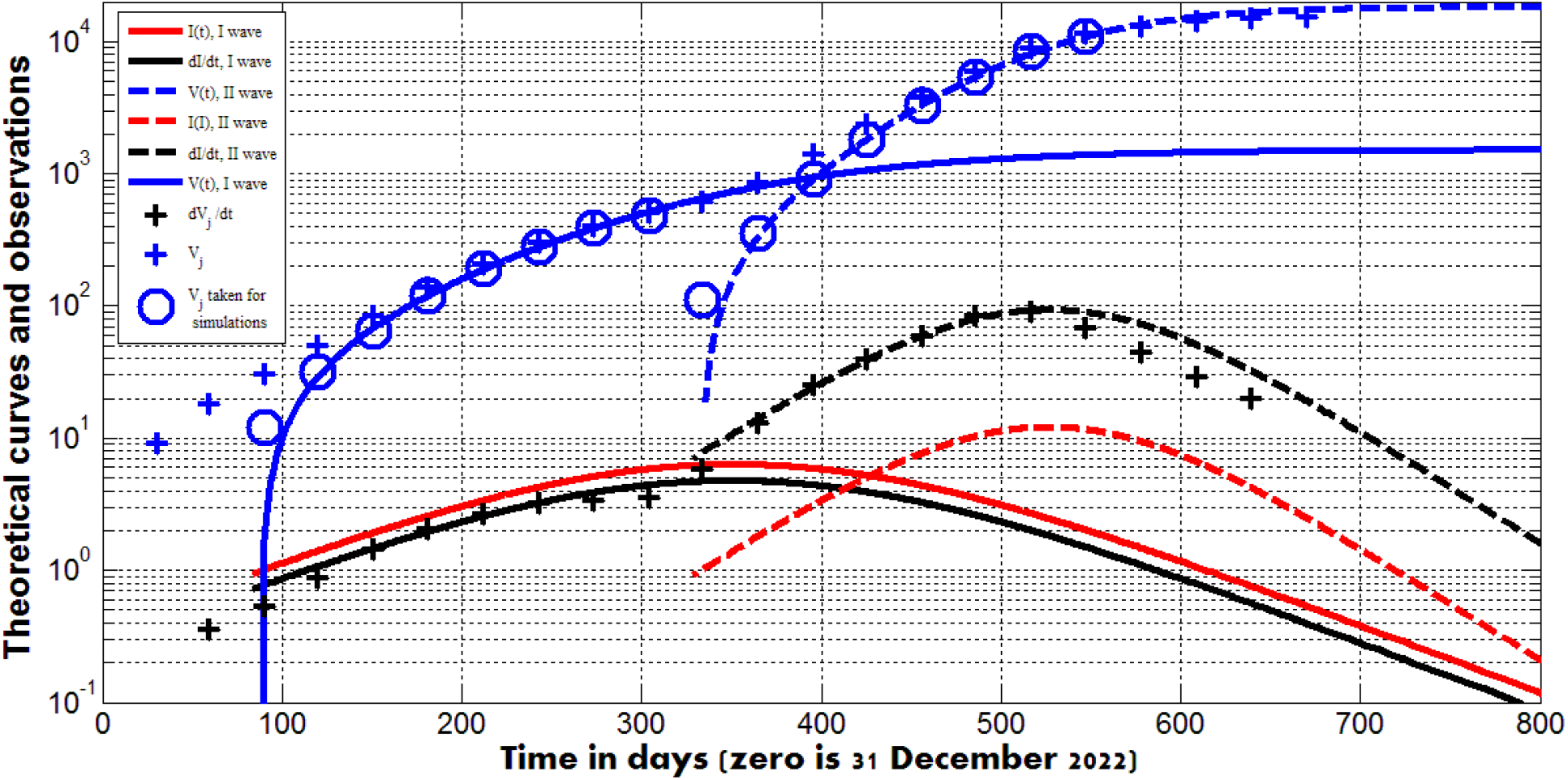
Results of simulations of the first (solid lines) and the second (dashed lines) waves and observations (markers).

## 5. Conclusions

To simulate hidden epidemic dynamics connected with asymptomatic and unregistered patients, a new general SIR model was proposed containing 5 unknown functions. For some cases, the analytical solutions of the set of 5 differential equations were found which allow simplifying the parameter identification procedure. Two waves of the pertussis epidemic in England in 2023 and 2024 were simulated for the case of zero hidden cases. Observations of accumulated visible numbers of cases during four months revealed rather high accuracy of predictions. If trend will not change, the monthly figure of 9 new pertussis cases (as it was in January-February 2023) can be achieved only in May 2025. The proposed approach can be recommended both for preliminary simulations of different epidemics (supposing zero hidden cases) and for further research using presented analytical solutions or numerical integration of differential equations.

### Clarification point

No humans or human data was used during this study

## Data availability

All data generated or analyzed during this study are included in this text.

## Conflict of interest

The author declares no conflict of interests.

## Acknowledgements

The author is grateful to James Robinson, Robin Thompson, Matt Keeling, Paul Brown, and Oleksii Rodionov for their support and providing very useful information. This paper was written with the support of the INI-LMS Solidarity Programme at the University of Warwick, UK.

## Notes

### Competing Interest Statement

The authors have declared no competing interest.

### Funding Statement

The study was supported by the INI-LMS Solidarity Programme at the University of Warwick, UK.

## References

1. https://edition.cnn.com/2020/11/02/europe/slovakia-mass-coronavirus-test-intl/index.html. Retrieved Novemver 23, 2024.

2. https://www.voanews.com/covid-19-pandemic/slovakias-second-round-coronavirus-tests-draws-large-crowds. Retrieved Novemver 23, 2024.

3. https://podillyanews.com/2020/12/17/u-shkolah-hmelnytskogo-provely-eksperyment-z-testuvannyam-na-covid-19/. Retrieved Novemver 23, 2024.

4. Schreiber, P.W., Scheier, T., Wolfensberger, A. et al. Parallel dynamics in the yield of universal SARS-CoV-2 admission screening and population incidence. Sci Rep 13, 7296 (2023). 10.1038/s41598-023-33824-6

5. Fowlkes A.L., Yoon S.K., Lutrick K., Gwynn L., Burns J., Grant L., Phillips A., Ellingson K., Ferraris M., LeClair M., et al. Effectiveness of 2-Dose BNT162b2 (Pfizer BioNTech) mRNA Vaccine in Preventing SARS-CoV-2 Infection Among Children Aged 5-11 Years and Adolescents Aged 12–15 Years - PROTECT Cohort, July 2021–February 2022. MMWR Morb. Mortal. Wkly. Rep. 2022;71:422–428. doi: 10.15585/mmwr.mm7111e1

6. Shang W, Kang L, Cao G, Wang Y, Gao P, Liu J, Liu M. Percentage of Asymptomatic Infections among SARS-CoV-2 Omicron Variant-Positive Individuals: A Systematic Review and Meta-Analysis. Vaccines (Basel). 2022 Jun 30;10(7):1049. doi: 10.3390/vaccines10071049. PMID: 35891214; PMCID: PMC9321237.

7. Rodger Craig et al. Asymptomatic Infection and Transmission of Pertussis in Households: A Systematic Review. Clin Infect Dis. 2020 Jan 1;70(1):152–161. doi: 10.1093/cid/ciz531.

8. Davies NG, Klepac P, Liu Y, Prem K, Jit M; CMMID COVID-19 working group; Eggo RM. Age-dependent effects in the transmission and control of COVID-19 epidemics. Nat Med. 2020 Aug;26(8):1205–1211. doi: 10.1038/s41591-020-0962-9. Epub 2020 Jun 16. PMID: 32546824.

9. Nesteruk, I. (2024): Trends of the COVID-19 dynamics in 2022 and 2023 vs. the population age, testing and vaccination levels. Front. Big Data 6:1355080. doi:10.3389/fdata.2023.1355080

10. Nesteruk, I., Keeling, M. (2023): Population age as a key factor in the COVID-19 pandemic dynamics. Preprint. Research Square. Posted November 30, 2023. 10.21203/rs.3.rs-3682693/v1

11. Nesteruk, I. (2021): Visible and real sizes of new COVID-19 pandemic waves in Ukraine. Innov Biosyst Bioeng., vol. 5, no. 2, pp. 85–96. DOI:10.20535/ibb.2021.5.2.230487 http://article/view/230487ibb.kpi.ua/

12. Nesteruk, I. (2021) Influence of Possible Natural and Artificial Collective Immunity on New COVID-19 Pandemic Waves in Ukraine and Israel. Exploratory Research and Hypothesis in Medicine; Published online: 11 November 2021. doi: 10.14218/ERHM.2021.00044.

13. Nesteruk, I. (2021) The real COVID-19 pandemic dynamics in Qatar in 2021: simulations, predictions and verifications of the SIR model. Semina: Ciências Exatas e Tecnológicas; 42:55–62. https://www.uel.br/revistas/uel/index.php/semexatas/article/view/43992

14. Kermack WO, McKendrick AG. A Contribution to the mathematical theory of epidemics. J Royal Stat Soc Ser A. 1927;115:700–21.

15. Weiss, H. (2013). The SIR model and the foundations of public health. MatMat 3, 1–17.

16. Daley DJ, Gani J (2005). Epidemic Modeling: An Introduction. New York: Cambridge University Press.

17. M. Keeling, P. Rohani, Modelling Infectious Diseases (Princeton Univ. Press, Princeton, NJ, 2008).

18. Cherniha, V. Davydovych, A Mathematical Model for the COVID-19 Outbreak and Its Applications, Symmetry 12 (2020) 990.

19. Alireza Mohammadi, Ievgen Meniailov, Kseniia Bazilevych, Sergey Yakovlev, Dmytro Chumachenko. Comparative study of linear regression and SIR models of COVID-19 propagation in Ukraine before vaccination. Radioelectronic and Computer Systems, 2021, Vol. 0, no. 3, pp. 5–18. DOI 10.32620/reks.2021.3.01

20. Nesteruk, I. COVID19 pandemic dynamics. Springer Nature, 2021, DOI: 10.1007/978-981-33-6416-5

21. Nesteruk, I. Detections and SIR simulations of the COVID-19 pandemic waves in Ukraine. Comput. Math. Biophys. 2021;9:46–65. 10.1515/cmb-2020-0117

22. Nesteruk, I. Improvement of the software for modeling the dynamics of epidemics and developing a user-friendly interface, Infectious Disease Modelling, Volume 8, Issue 3, 2023, Pages 806–821, ISSN 2468-0427, 10.1016/j.idm.2023.06.003.

23. Nesteruk I. Statistics based models for the dynamics of Chernivtsi children disease. Naukovi Visti NTUU KPI. 2017;5:26–34. DOI: 10.20535/1810-0546.2017.5.108577

24. Hethcote HW (2000). The mathematics of infectious diseases. SIAM Review. 42 (4): 599–653.

25. Nakamura, Gilberto M.; Cardoso, George C.; Martinez Alexandre S. (February 2020). Improved susceptible–infectious–susceptible epidemic equations based on uncertainties and autocorrelation functions. Royal Society Open Science. 7 (2): 191504.

26. Britton NF. Essential Mathematical biology, Springer (India) Pvt. Limited, 2004, 352 p.

27. Pesco P, Bergero P, Fabricius G, Hozbor D. Modelling the effect of changes in vaccine effectiveness and transmission contact rates on pertussis epidemiology. Epidemics. 2014 Jun;7:13–21. doi: 10.1016/j.epidem.2014.04.001.

28. Nesteruk, I. Endemic characteristics of SARS-CoV-2 infection. Sci Rep 13, 14841 (2023). 10.1038/s41598-023-41841-8

29. Confirmed cases of pertussis in England by month - GOV.UK (www.gov.uk)

30. Nesteruk, I. (2024): Mathematical simulations of the pertussis epidemic in England. International Workshop ProfIT AI 2024, Cambridge, MA, USA, 25-27.09.2024. https://ceur-ws.org/Vol-3777/paper13.pdf

31. Draper NR, Smith H. Applied regression analysis. 3rd ed. John Wiley; 1998.

